# Survival rate of dental implants installed by residents attending an implantology Program in Brazil: a 52-month retrospective analysis

**DOI:** 10.1101/2022.09.04.22279582

**Authors:** Myungjin Kang, Henrique Smanio Neto, André Antonio Pelegrine, Cecilia Pedroso Turssi, Juliana Trindade Clemente-Napimoga, Marcelo Henrique Napimoga

**Affiliations:** Faculdade São Leopoldo Mandic, Instituto São Leopoldo Mandic, Implantology, Campinas, SP, Brazil; Faculdade São Leopoldo Mandic, Instituto São Leopoldo Mandic, Restorative Dentistry, Campinas, SP, Brazil; Faculdade São Leopoldo Mandic, Instituto São Leopoldo Mandic, Laboratory of Neuro-Immune Interface of Pain Research, Campinas, SP, Brazil

**Keywords:** Implantology, dentistry, dental implant, implant survival, education

## Abstract

The aim of this study was to identify any associations between predictor variables, mainly risk factors and dental implant outcome. Dental records were reviewed from January 1, 2018, to December 31, 2020. The inclusion criteria was all implant surgery made using Intraoss brand. Data collected from the patients’ medical charts included: implant loss, gender, diabetes, smoking, continuous use of medication, type of implant connection system, implant position (maxilla or mandible), previous bone grafting and type of prosthetic provisioning (temporary prosthesis, immediate prosthesis or permanent prosthesis). It was evaluated the cumulative survival rate of 1,164 dental implants made by residents attending an implantology residency in a university setting. One thousand forty-eight dental implants were placed on 471 patients seen by residents. The cumulative survival rate was 2.5%. Furthemore, the association of implant losses to the variables tested using the chi-square and G tests showed no statistically significant association. Based on Kaplan-Meier curve analysis, with a 95% confidence interval up to 52 months of implant placement, it revealed that the overall survival rate was 90.5%. Therefore, this study showed high survival rates of implants installed by residents of implantology at Faculdade São Leopoldo Mandic using Intraoss implants. The implant failure was not correlated with any of the variables tested.

## Introduction

The use of dental implants is considered one of the prominent scientific breakthrough and predictable treatment options to restore partially and totally edentulous patients. Thus, the large-scale use of dental implants, which demonstrate predictable long-term results from a functional, aesthetic and peri-implant health point of view, has high survival rates well demonstrated in the literature [1]. Recent studies reported 86% to 98% survival rates for dental implants after 5 years of follow-up [2,3] and around 90% even after 10 years of follow-up [4,5].

Implant-related complications have been categorized into two main types: biological and technical. Among the biological complications, some of the patient-related risk factors include: smoking and systemic diseases such as uncontrolled diabetes mellitus (DM); and periodontitis; which are all characterized as patient-related risk factors for implant failure [6,7]. From a technical point of view, clinical training in implant dentistry provides to graduate dental students advanced skills. Although many studies report the success of implant rehabilitations, there is limited literature on the survival of implants performed by residents students. A recent study evaluated the survival rates of implants and prostheses placed by undergraduate students in a dental hospital. The study was a retrospective university/hospital based study and included patients visiting the dental hospital. Of the 86,000 patients who visited Saveetha Dental College, a total of 79 patients were enrolled in the study according to the inclusion criteria of patients who had undergone implant surgery by undergraduate students. The survival rate from implants placed was 92.4% [8]. Another study based in the rehabilitation of patients with implants at the University of Alberta (Canada) by undergraduate students, evaluated 289 implants in 189 patients, with only 1 loss. Therefore, a high survival rate of 99.7% was verified [9].

The Brazilian implant industry (Intraoss, Itaquaquecetuba, Brazil) has different implant abutment connections such as external connection, internal connection and tapered connection, all of them fabricated from grade 5 medical titanium. Previous reports that have evaluated implants of this company demonstrated an effective bacterial seal at the implant/abutment interface between an external hexagon and a tapered connection system [10]. Besides, the dental implant surface treatment positively affected the early events of the interaction between titanium and osteoblastic cells [11].

Hence, the aim of this single-center study was to investigate retrospectively the survival rate of Intraoss implants performed by implantology resident students in Brazil.

## Material and Methods

### Ethics

The research project was approved by the Research Ethics Committee of Faculdade São Leopoldo Mandic, registration number # 49980221.7.0000.5374. All patients included in this study provided informed consent prior to implant treatment.

### Study subjects

The present study is a single-arm, retrospective observational study based on the dental implants performed by resident students in Implantology at São Leopoldo Mandic (Brazil) between January 1, 2018, and December 31, 2020, based on the data available in the medical records of patients. This observational study was conducted according to the guidelines of Strengthening the reporting of observational studies in epidemiology (STROBE).

### Inclusion criteria

Patients between 17 and 82 years of age whose records indicated that they received Intraoss dental implants system between January 1, 2018, and December 31, 2020.

### Exclusion criteria

The exclusion criteria were patients whose records indicated that they received another dental implant system during the analyzed period.

### Data Collection and Analyses

Data collected from the patients’ medical charts were submitted to descriptive and inferential analyses, using chi-square and G tests, to investigate the association between implant loss and gender, diabetes, smoking, continuous use of medication, type of implant connection system, implant site (maxilla or mandible), previous bone grafting and type of prosthetic provisioning (temporary prosthesis, immediate prosthesis or permanent prosthesis). The implant survival rate was estimated using the Kaplan-Meier curve. The level of significance was set at 5% and statistical calculations was performed using SPSS 23 (SPSS INC., Chicago, IL, USA) and BioEstat 5.0 (Fundação Mamirauá, Belém, PA, Brazil).

## Results

During the 2018 – 2020 period, residents in Implantology at São Leopoldo Mandic performed a total of 3,875 implants. According to the inclusion criteria, a total of 1,164 dental implants, which had been installed in the oral cavity of 742 patients were included in the study. Of the total number of patients included in this study 254 (34.2%) were male and 486 (65.5%) were female. For two (0.3%) patients, information regarding gender was non-existent. The age of the patients ranged from 17 to 82 years old (average: 55.1 ± 11.5 years). Only one of the 742 patients had no information regarding age.

Fifty-four (7.3%) out of the 742 patients had diabetes, with 26 (3.5%) being male and 28 (3.8%) female, while 680 (91.6%) had no diabetes and for 8 patients this information was unavailable.

Smoking was identified in 96 (12.9%) of the 742 patients, of whom 40 (5.4%) were men and 56 (7.5%) were women. Non-smokers summed 643 (86.7%) patients and three others had no information about smoking. Sixteen (2.2%) patients were both smokers and diabetics.

Of the 742 patients, 354 (47.7%) were on continuous medication, of which 109 (31.9%) were men, 244 (32.9%) were women and one had gender uninformed. Non-users of medication totaled 385 (51.9%) patients and other three did not have information on this aspect.

In the oral cavity of the 742 patients, 1,164 implants were installed, indicating an average of 1.6 implants per patient. The maximum number was six implants in the same patient. Of the 1,164 implants installed, 907 (77.9%) were tapered connection system, 174 (14.9%) were external connection and 80 (6.9%) were internal connection. For three implants the system was unknown.

Of the 1,164 implants, 605 (52.0%) were installed in the maxilla, 530 (45.5%) in the mandible, and for 27 of them the location was not indicated. Bone grafting procedures preceded the installation of 278 (23.9%) of the 1,164 implants, while for the remaining 886 there was no grafting or this information was non-existent (for three implants).

The mean time of implant installation prior to the data collection was 14.2 months (± 10.2 months), with the shortest time being one month and the longest 52 months. Figure 1 is a histogram showing the number of implants that had been installed within each 6-month period. More than half (601) of the 1,164 implants, corresponding to 51.6%, had been installed up to 12 months prior to the data collection, while 992 (85.2%) had been installed up to 24 months previously. For two implants, there was no information on the installation time.

**Figure 1:**
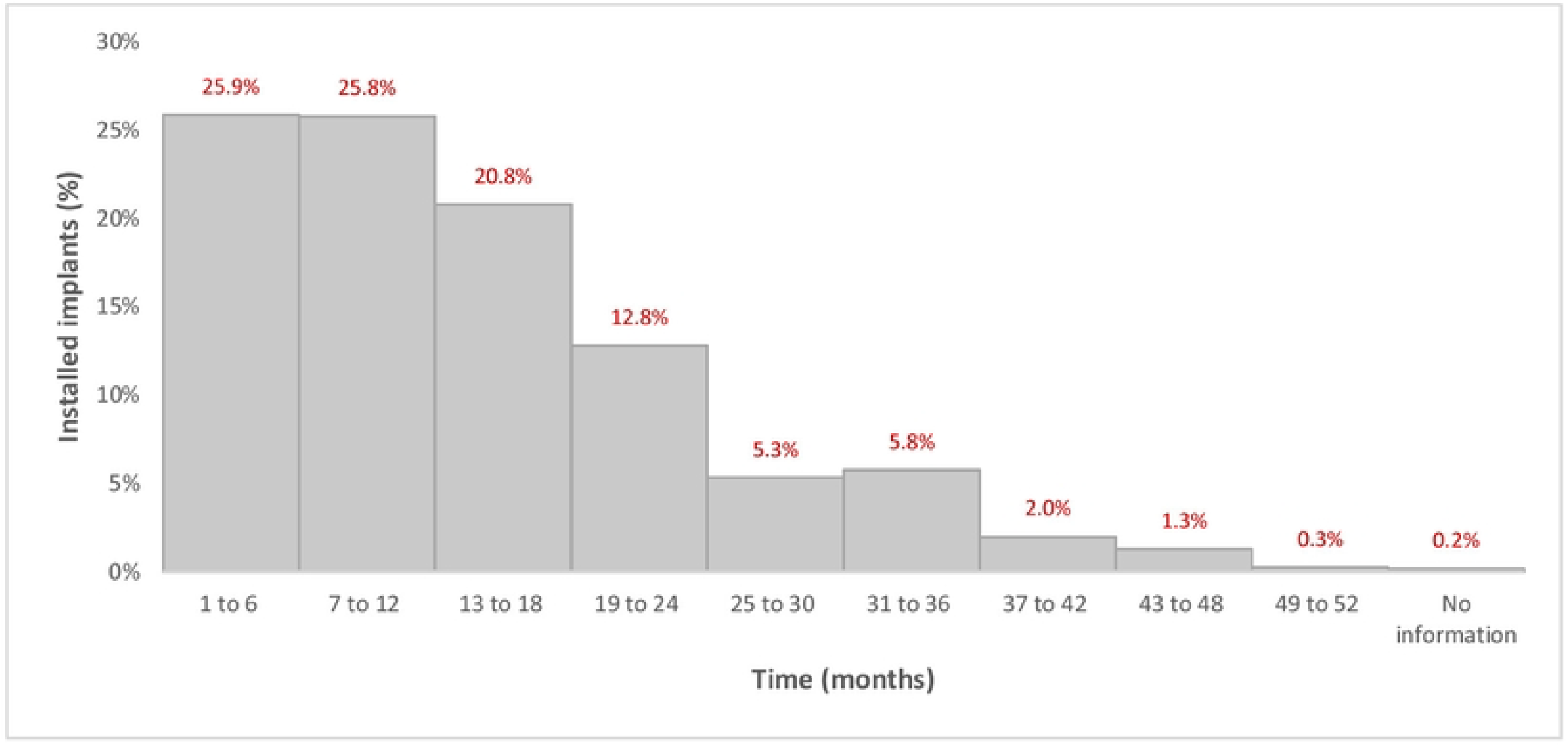
Histogram of the installation time of the evaluated implants.

Among the 1,164 implants, 385 (33.1%) received a temporary prosthesis, 257 (22.1%) received an immediate prosthesis and 364 (31.3%) received a permanent prosthesis. Provisionalization information was absent for 158 implants.

Of the 1,164 implants installed, 29 (2.5%) failed. Investigating the association of implant losses with valid responses (excluding cases with no information) there was no statistically significant association with gender, diabetes, smoking, continuous use of medication, type of implant system, implant placement site, previous bone grafting and type of prosthetic provisioning (Table 1).

**Table 1.** – Absolute and relative frequencies (%) of implant loss according to sex, diabetes, smoking, continuous use of medications, type of implant system, location of installation, previous bone grafting and type of prosthetic provisioning.

| Independent Variable | Implant loss |  | p-value |
| --- | --- | --- | --- |
|  | Y | N |  |
| Gender |  |  |  |
| Male | 11 (2.7%) | 398 (97.3%) | 0,755* |
| Female | 18 (2.4%) | 735 (97.6%) |  |
|  | 29 | 1.133 |  |
| Diabetes |  |  |  |
| Y | 2 (2.5%) | 78 (97.5%) | 0,966** |
| N | 26 (2.4%) | 1.047 (97.6%) |  |
|  | 28 | 1.125 |  |
| Smoking |  |  |  |
| Y | 6 (3.8%) | 154 (96.2%) | 0,274* |
| N | 23 (2.3%) | 978 (97.7%) |  |
|  | 29 | 1.132 |  |
| Continuous use of medications |  |  |  |
| Y | 12 (2.1%) | 555 (97.9%) | 0,416* |
| N | 17 (2.9%) | 577 (97.1%) |  |
|  | 29 | 1.132 |  |
| Implant System |  |  |  |
| Tapered connection | 21 (2.3%) | 886 (97.7%) | 0,704** |
| External connection | 5 (2.9%) | 169 (97.1%) |  |
| Internal connection | 1 (1.3%) | 79 (98.7%) |  |
|  | 27 | 1.134 |  |
| Implant placement site |  |  |  |
| Maxila | 11 (1.8%) | 594 (98.2%) | 0,132* |
| Mandible | 17 (3.2%) | 513 (96.8%) |  |
|  | 28 | 1.107 |  |
| Previous bone grafting |  |  |  |
| Y | 7 (2.5%) | 271 (97.5%) | 0,895* |
| N | 21 (2.4%) | 862 (97.6%) |  |
|  | 28 | 1.133 |  |
| Prosthesis |  |  |  |
| Temporary prosthesis | 8 (2.1%) | 377 (97.9%) | 0,543** |
| Immediate prosthesis | 5 (1.9%) | 252 (98.1%) |  |
| Definitive prosthesis | 10 (2.7%) | 354 (97.3%) |  |
|  | 23 | 983 |  |

For each independent variable, cases without information were disregarded for applying the analyses. \* = p-value of the chi-square test; \*\* = p value referring to the G test

Among the 29 implants that failed, in one of them no information about the installation time was present. The same lack of information occurred within an implant that did not fail. Therefore, of the 1,164 implants installed, 1,162 implants were considered for estimating the survival rate. Figure 2 shows the Kaplan-Meier curve, with a 95% confidence interval up to 52 months after implant placement, and reveals that the overall survival rate was 90.5%. Table 2 indicates the survival rates in the other time intervals and presents the confidence intervals (95%).

**Table 2.**
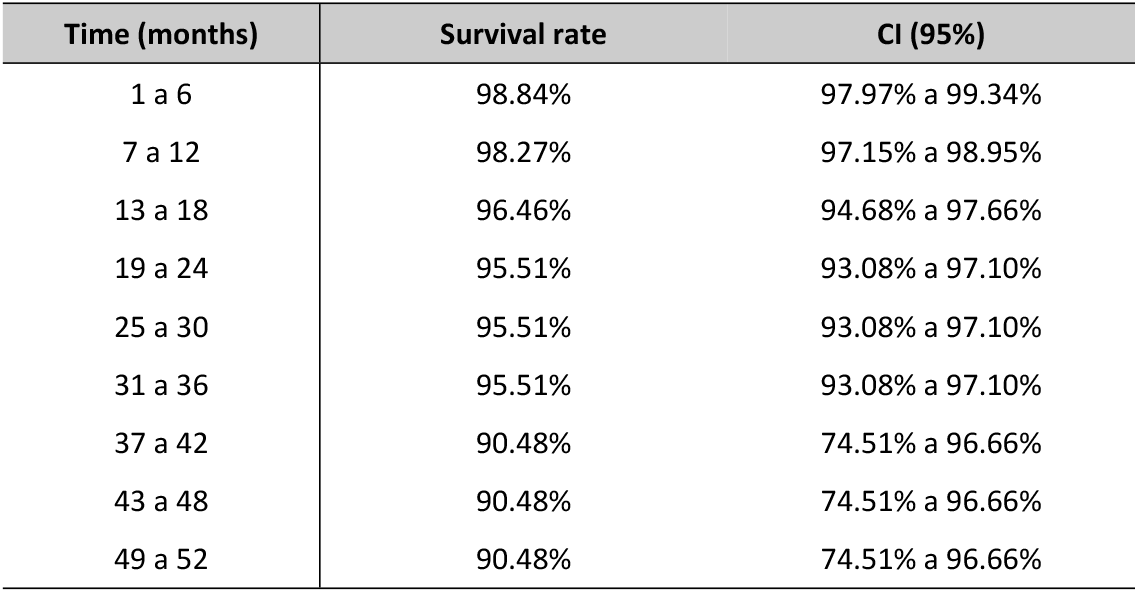
Survival rate and confidence interval (95% CI) according to implant placement period.

**Figure 2:**
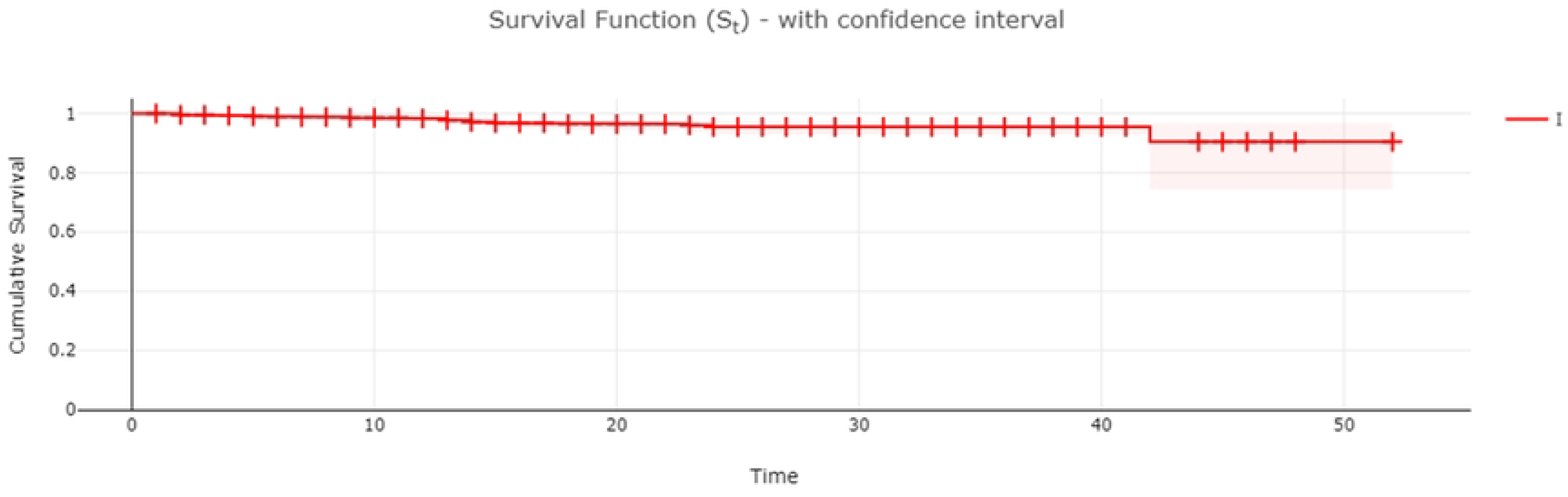
Kaplan Meier curve for the event of implant loss in the sample evaluated.

## Discussion

This study showed the high survival rate of Intraoss implants system made by residents in Implantology Program at São Leopoldo Mandic (Brazil), up to 52 months and none of the independent variables tested positively correlated with the implant failure. This retrospective study included 742 patients who received a total of 1,164 implants showing a cumulative failure rate of 2.5%. Based on Kaplan-Meier curve analysis, with a 95% confidence interval up to 52 months of implant placement, revealed that the overall survival rate at this time was 90.5%.

Some individual biological factors are known to potentially impair implant prognosis. A meta-analyses based on implant- and patient-related data showed a significant increase in the relative risk of implant failure in patients who smoked >20 cigarettes per day compared with non-smokers [12]. In the present study we did not observe a positive association of smoking and dental implant failure, however, we did not have the information of how many cigarettes were used per day by each patient, which might biased the results. Importantly, it cannot be ruled out that the lack of association between smoking and failure can be partly attributed to the time since implant placement in our study was up to 52 months. In addition, when working with a dichotomous assessment (yes vs. no for smoking) it is difficult to show effects that are known to be more expressive, thus, if number of cigarettes/packs categorizes it may favors finding an association.

The long-term hyperglycemia of diabetes usually leads to failure, damage, and/or dysfunction of many tissues and organs mainly due to the correlation between glycemic control and the development of microvascular and macro-vascular complications [13]. Previous results already demonstrated that diabetic patients presented a statistically significant higher risk of dental implant failure and higher marginal bone loss than non-diabetic patients, mainly type 1 diabetes [14]. Our results have not demonstrated any statistical significance regarding diabetes and dental implant failure, however the data about the presence or absence of diabetes was based only on patient’ s information since no laboratorial test was made, and thus this data may be carefully analyzed. However, another study verified no association between implant loss and different variables such as bone augmentation, time of implant placement, diabetes and smoking, corroborating our results [15]. Besides, in a previous study several parameters similar to those evaluated here did not yield any significant association with implant failures [16].

Clinical training in implant dentistry for graduate students contributes to the development of advanced skills in dental students. Data from a study performed by undergraduate students who carried out the rehabilitation of patients with implants at the University of Alberta (Canada), who installed 289 implants in 189 patients, with only 1 loss occurred, and therefore, a survival rate of 99.7% was achieved [9]. On the other hand, On the other hand, the influence of surgeons’ dental/implant education and its relevance to treatment outcome on implant failure rates has been shown to be important. In a previous study the rates obtained at International University of Catalonia (Barcelona, Spain) resulted in overall cumulative rates of 4.9% and 10.8% at the implant and patient levels, respectively, over a 7-year period [15]. The success rate for Harvard School of Dental Medicine periodontology residents was 96.48% during the 4-year study period [17]. Nonetheless, an interesting study analyzed the implant outcomes and the clinical training at Louisiana State University Health Science Centera (USA), showing that the advanced group (94.2%) had the best implant outcomes followed by the intermediate group (89.38%) and beginner group (88.6%) clearly demonstrating that increased clinician training improves clinical outcomes [16].

Moreover, an interesting analysis demonstrated that clinicians’ age and years of experience as dentists or as specialist were not found to be predictors to early implant failure rate however, the number of implants placed during the postgraduate training was found to be significantly predicting early failure rate of implants [18]. In the present study, of the 1,164 implants installed by residents in implantology, 29 (2.5%) failed up to 52 months after implant placement. It reveals an overall survival rate of 90.5% at this time. Thus, the present data is very close to published data by several universities worldwide during the residency training.

Important to point out that a limitation of the study was the retrospective timeline of up to 52 months and the fact that the implants included had been present for variable times as well as almost 85% of the installed implants had up to 2 years. The lower number of implants installed more than 3 years induced the largest confidence interval observed (Table 2).

The present study showed high survival rates made by residents of implantology at Faculdade São Leopoldo Mandic using Intraoss dental implants, and the implant failure had no association with any of the variables tested.

## Data Availability

Data are available from the São Leopoldo Mandic Institutional Data Access (contact via)

## Acknowledgements

This work was supported in part by the Conselho Nacional de Desenvolvimento Científico e Tecnológico (CNPq) - Research Productivity Fellowship to MHN and JTCN; Coordenação de Aperfeiçoamento de Pessoal de Nível Superior (CAPES) - #001.

## Author Contributions section

M.K., T. C.-N., A.A.P., H.S.N., M. H. N. designed research

M.K. performed research

C.P.T. performed the statistical analysis

M.K., H.S.N., A.A.P., C.P.T., J.T.C-N., M.H.N. wrote the paper

All authors read and approved the final version of the manuscript.

